# Temporal Clustering of Acute Neurological Disorders: Testing the Clinical Impression of Diagnostic “Theme Shifts”

**DOI:** 10.64898/2026.08.28.26361586

**Authors:** LAL Härtel, A Jaeger, FA Riethues, J von Itter, H Lee, S Hause, S Meuth, A Schmidt-Pogoda

## Abstract

**Background:** On-call clinicians frequently report the anecdotal impression of “theme shifts” during which specific acute neurological diagnoses appear to cluster. Whether such clustering reflects a statistically true and reproducible phenomenon has not been systematically investigated; the present paper examines seasonality and temporal clustering within six different acute neurological conditions.

**Methods:** In this retrospective, single-center cohort study, we identified all patients admitted to a tertiary neurological department between July 2016 and June 2026 with acute unilateral vestibulopathy, cerebral artery dissection, generalized epileptic seizures, primary intracerebral hemorrhage, peripheral facial nerve palsy, or transient global amnesia (TGA) (n = 2,140). Monthly and seasonal distributions were assessed using chi-squared goodness-of-fit and cosinor analysis. Short-term temporal clustering was tested by Monte Carlo permutation across time windows from 24 hours to 90 days, and endogenous cluster dynamics were characterized using Hawkes self-exciting point process modeling.

**Results:** Admissions for generalized epileptic seizures showed a statistically significant deviation from a uniform monthly distribution with a winter distribution (p<0.001 and q = 0.002), and a significant temporal clustering across time windows from 72 hours to 90 days (all q < 0.05). Peripheral facial nerve palsy presented significant clustering at the 90-day window (q = 0.029) and TGA at 60-day time window (q = 0.041) without seasonality; the diagnostic groups of acute unilateral vestibulopathy, cerebral artery dissection and primary intracerebral hemorrhage showed neither seasonality nor clustering after correction for multiple comparison. No diagnostic group showed clustering within a 24-hour window, statistically significant self-excitation in Hawkes process modelling, or a significant linear trend in monthly case counts over the study period.

**Conclusion:** The anecdotal impression of diagnostic “theme shifts” among on-call neurologists appears to have a measurable basis, although clustering is confined to specific conditions and rather on a time scale of weeks to months. Generalized epileptic seizures were the only diagnostic group that uniquely combined seasonality with temporal clustering, suggesting a shared trigger, while facial palsy and TGA showed episodic, yet non-seasonal clustering.

## Introduction

Clinicians across generations have reported the impression of “theme weekends” or “theme days” during on-call shifts, when specific acute neurological diseases appear to cluster, e.g. shifts remembered as yet another “bleeding weekend”. Such observations may be partly attributed to cognitive bias, yet they still raise the fascinating question of whether certain acute neurological conditions tend to occur in temporal clusters or follow seasonal patterns of incidence.

Seasonality and temporal clustering have been described for several acute neurological conditions ^1–7^. Subarachnoid hemorrhage, primary intracerebral hemorrhage, and ischemic stroke have all been linked to seasonal variation ^1, 2, 4, 5, 8–10^, with a predilection for colder months ^2, 5, 9^, potentially mediated by blood pressure fluctuations ^11^, hypercoagulability ^12–14^, and increased rates of respiratory infections in winter ^15, 16^. Peripheral facial nerve palsy has been associated with seasonal variation, with several studies reporting higher incidence during colder months, potentially related to environmental conditions and viral reactivation ^6, 7, 17–19^. Epileptic seizures have also been linked to infectious and environmental triggers, although evidence for a consistent seasonal distribution is less robust ^20^.

Dissections of the cervical and intracranial arteries supplying the brain are a rare but clinically significant origin of stroke, particularly in younger patient cohorts ^21, 22^. Structural vascular vulnerability, hemodynamic stress, and possibly inflammatory or infectious triggers have been proposed as contributing mechanisms ^22, 23^. For the more common cervical artery dissections a predominance in the colder months has been reported in several studies ^24–27^, suggesting a potential link to upper respiratory tract infections inducing local inflammation or increased intrathoracic pressure resulting from severe coughing ^27, 28^.

To our knowledge, no prior study has systematically analyzed the temporal and seasonal distribution of acute neurological diseases, nor compared these patterns. We therefore conducted a retrospective analysis of all patients admitted to the Department of Neurology at the University Hospital Münster between July 1st, 2016, and June 30th, 2026, with one of the following diagnoses: cerebral artery dissection, primary intracerebral hemorrhage (ICH), generalized epileptic seizure, idiopathic peripheral facial palsy (Bell’s palsy), vestibular neuronitis, and transient global amnesia (TGA). For each condition individually, the primary objective was to determine whether admissions show significant seasonal variation or temporal clustering in incidence. In addition to that, examination of endogenous cluster dynamics was conducted to quantify the mathematical dynamics and intensity of these transient clusters.

This approach may help distinguish perceived from measurable temporal patterns and provide a basis for further investigation of potential shared triggers.

## Methods

### Study Design and Setting

This was a retrospective, single-center cohort study conducted at the University Hospital Münster, a tertiary care academic medical center and comprehensive stroke center in northwestern Germany. The study was approved by the local ethics committee (2026-650-f-S). Given the retrospective, non-interventional design using routinely collected clinical data, informed consent was waived.

### Patient Selection

All patients admitted between July 1^st^, 2016, and June 30^th^, 2026, with a primary discharge diagnosis of the following acute neurological diseases were identified from the hospital information system (HIS) using the following ICD-10-GM codes: I67.0 (dissection of cerebral arteries), I72.0, I72.5, I72.6, I72.8, and I72.9 (dissection and aneurysm of cerebral arteries). Furthermore, we identified admissions from the hospital information system (HIS) with the following ICD-10-GM codes: G40.6 (generalized epileptic seizure), G45.42, G45.43, and G45.49 (transient global amnesia), G51.0 (peripheral facial nerve palsy), H81.2 (acute unilateral vestibulopathy), I61.0, I61.1, I61.2, I61.3, I61.4, I61.5, I61.6, I61.8, and I61.9 (intracerebral hemorrhage), R56.8 (other and unspecified convulsions).

Patients with incomplete admission date records were excluded, the primary discharge diagnosis code was confirmed through medical transcripts. In cases where the exact onset of symptoms was unavailable, the date of hospital admission was used as a proxy for the date of the neurological event, as is standard practice in retrospective registry studies. This approximation is discussed as a study limitation.

### Data Variables

The following variables were extracted for each case: date of admission, ICD-10 discharge diagnosis.

### Temporal and Seasonal Variables

From the admission date we derived the calendar month (1-12), the meteorological season (Winter, Spring, Summer, Fall) and the calendar year for each admission. The seasonal definition followed standard meteorological conventions applied in climate and epidemiological research, with Winter lasting from December through February, Spring from March through May, Summer from June through August, and Autumn from September through November. The admission data also included the exact admission time for analysis of smaller time clusters.

### Statistical Analysis

Based on distributional assessment, continuous variables are reported as mean with standard deviation or median with interquartile range. Categorical variables are reported as absolute counts and percentages. All analyses were performed using R (version 4.5.1, R Foundation for Statistical Computing, Vienna, Austria). Each of the six diagnostic groups were analyzed in parallel within an identical analytical framework, and thus each constituting a family of simultaneous hypothesis tests. To account for a possible false discovery rate (the expected proportion of false positives among the hypotheses declared as significant), p-values were adjusted for multiple comparisons using the Benjamini-Hochberg procedure ^29^. This method was chosen due to the exploratory and hypothesis-generating character of the present study rather than confirmatory. The adjustment was applied within, rather than across, analysis families (secular trend, deviation from a uniform monthly distribution, annual rhythmicity, short-term temporal clustering, and endogenous self-excitation). Within the permutation family (Monte Carlo simulation), the thirty-six tests (six diagnostic groups x six time windows) were positively dependent, the Benjamini-Hochberg procedure allowed to retain control of false discovery rates in this form of positive dependence ^30^. Both unadjusted p-values and adjusted q-values are reported throughout. Statistical significance was assessed on Benjamini-Hochberg adjusted q-values, with q < 0.05 considered significant; unadjusted p-values are reported alongside for transparency.

#### Analysis of frequency and seasonality

Assessment of temporal trends in monthly case counts was collected using linear regression with the month as the independent variable, and for evaluation whether the monthly distribution of cases differed significantly from uniform distribution, we performed a chi-squared goodness-of-fit test.

For each of the different acute neurological disease seasonality was examined by using cosinor analysis, modeled as a linear regression with cosine and sine terms representing the annual (12-month) and quarterly (3-month) periodicity, which allowed for quantification of the amplitude and acrophase of any rhythmic component. The null hypothesis of no seasonality (zero amplitude at both periods) was tested by the F-test comparing this model against a model containing the linear time trend alone, so that the test reflects rhythmic variation with any secular trend already removed.

#### Temporal clustering and Monte Carlo simulations

To examine the clinical hypothesis that the chosen clinical pictures exhibit non-random, short-term clusters, we performed a Monte Carlo simulation. Whilst retaining the global distribution characteristics, a Monte Carlo simulation was used to generate a permutation null distribution against which the empirically observed number of close event pairs was compared.

The empirically observed number of event pairs separated by no more than each predefined time window was tested against the permutation null distribution to determine whether close event pairs occurred more frequently than. The null distribution was generated by random labelling: for each diagnostic group with n events, n admission timestamps were drawn at random without replacement from the pooled set of all admission timestamps across the six diagnostic groups (999 permutations), which allowed for preserving the overall structure of acute neurological admissions, including administrative or day-of-week patterns. The time windows chosen were 90 days, 60 days, 30 days, 7 days, 72 hours and 24 hours to achieve a higher probability of detection of temporal clustering in the different chosen diseases. The test statistic was the number of event pairs separated by no more than a given time window; p-values are one-sided permutation p-values.

#### Endogenous cluster dynamics (Hawkes Process modelling)

For further characterization of any identified temporal clusters, a self-exciting point process model was applied (Hawkes Process). Assuming that each observed event temporarily increases the likelihood of subsequent events occurring in close temporal proximity, this modelling approach captures the clustering dynamics inherent to the data. At any given time point, the underlying event rate is modelled as the sum of two elements: a time-varying background rate, where long-term seasonal fluctuations are integrated, that were previously determined by the seasonal cosinor analysis, and a short-term excitation element that represents the transient increase in event probability following each preceding event. The elevation is only transient as it decays exponentially over time, and by that diminishing the influence of any single event progressively. The parameters of the time-varying background rate were estimated in the preceding cosinor analysis and subsequently held fixed, whereas the excitation magnitude α and decay rate β were estimated by maximum likelihood. The conditional intensity was λ(t) = μ(t) + Σ α·exp(−β(t−tᵢ)) over all earlier events tᵢ, where μ(t) is the fitted cosinor background rate from the seasonality analysis; the branching ratio is α/β. Parameters were estimated by maximum likelihood using Nelder-Mead optimization from multiple starting values. Significance of self-excitation (H₀: α = 0) was assessed by a bootstrap-calibrated likelihood-ratio test, because α = 0 lies on the boundary of the parameter space, which makes the standard chi-squared reference distribution anti-conservative. The bootstrap p-value is the proportion of simulated likelihood-ratio statistics (generated from an inhomogeneous Poisson process with the fitted seasonal background) at least as extreme as the observed statistic.

The full mathematical specification of the model, including the conditional intensity function and parameter definitions, is provided within the full R script, which is available upon request.

## Results

### Study Population

Across the six diagnostic groups a total of 2,140 admissions were included: Acute unilateral vestibulopathy (n=112), dissection of cerebral arteries (n=177), generalized epileptic seizures (n=580), non-traumatic intracerebral hemorrhage (n=864), peripheral facial palsy (n=214) and transient global amnesia (TGA, n=193), events per year ranged from 11.2 to 86.4 per diagnostic group. Baseline characteristics are presented in Table 1.

**Table 1.** Baseline characteristics of the study population by diagnostic group.

| Entity | Acute unilateral vestibulopathy | Dissection of cerebral arteries | Generalized epileptic seizures | ICH<sup>a</sup> | Peripheral facial nerve palsy | TGA<sup>b</sup> |
| --- | --- | --- | --- | --- | --- | --- |
| n | 112 | 177 | 580 | 864 | 214 | 193 |
| First event | 2016-09-26 | 2016-07-06 | 2016-07-10 | 2016-07-02 | 2016-07-12 | 2016-08-03 |
| Last event | 2026-05-23 | 2026-04-24 | 2026-06-11 | 2026-06-17 | 2026-06-13 | 2026-05-05 |
| Study period (years) | 10.00 | 10.00 | 10.00 | 10.00 | 10.00 | 10.00 |
| Events per year | 11.2 | 17.7 | 58.0 | 86.4 | 21.4 | 19.3 |
| Median IEI<sup>c</sup> (days) | 24.59 | 15.17 | 4.00 | 2.92 | 11.84 | 12.85 |
| Q1 IEI (days) | 9.46 | 6.00 | 1.65 | 1.22 | 5.14 | 5.08 |
| Q3 IEI (days) | 47.22 | 29.21 | 8.27 | 5.80 | 21.42 | 26.27 |

<sup>a</sup> ICH = intracerebral hemorrhage; <sup>b</sup> TGA = transient global amnesia; <sup>c</sup> IEI = inter-event interval

### Temporal Trend

The absence of a statistically significant temporal trend, observed by linear regression, in monthly case counts for any of the six diagnostic groups over the observation period (all six diseases p > 0.05; Table 2, Figure 1) suggests that the findings are not systematically confounded by changes in referral patterns, diagnostic awareness, or coding practices in the observed time period.

**Figure 1.**
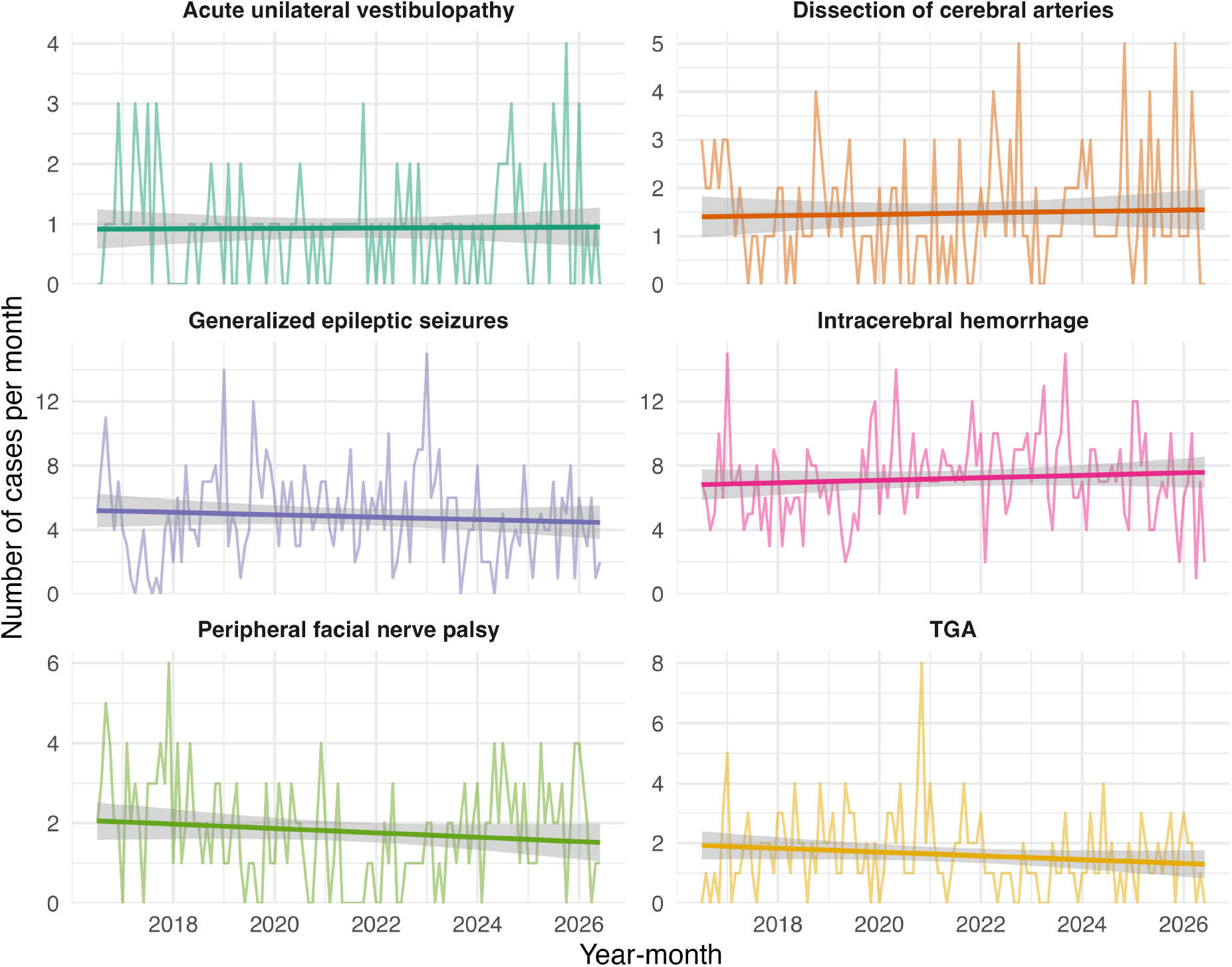
Monthly case counts over the 10-year observation period from 2016 to 2026 for each of the six diagnostic groups, with linear regression trend line. No diagnostic group showed a statistically significant temporal trend (all p > 0.05; see Table 2).

**Table 2.**
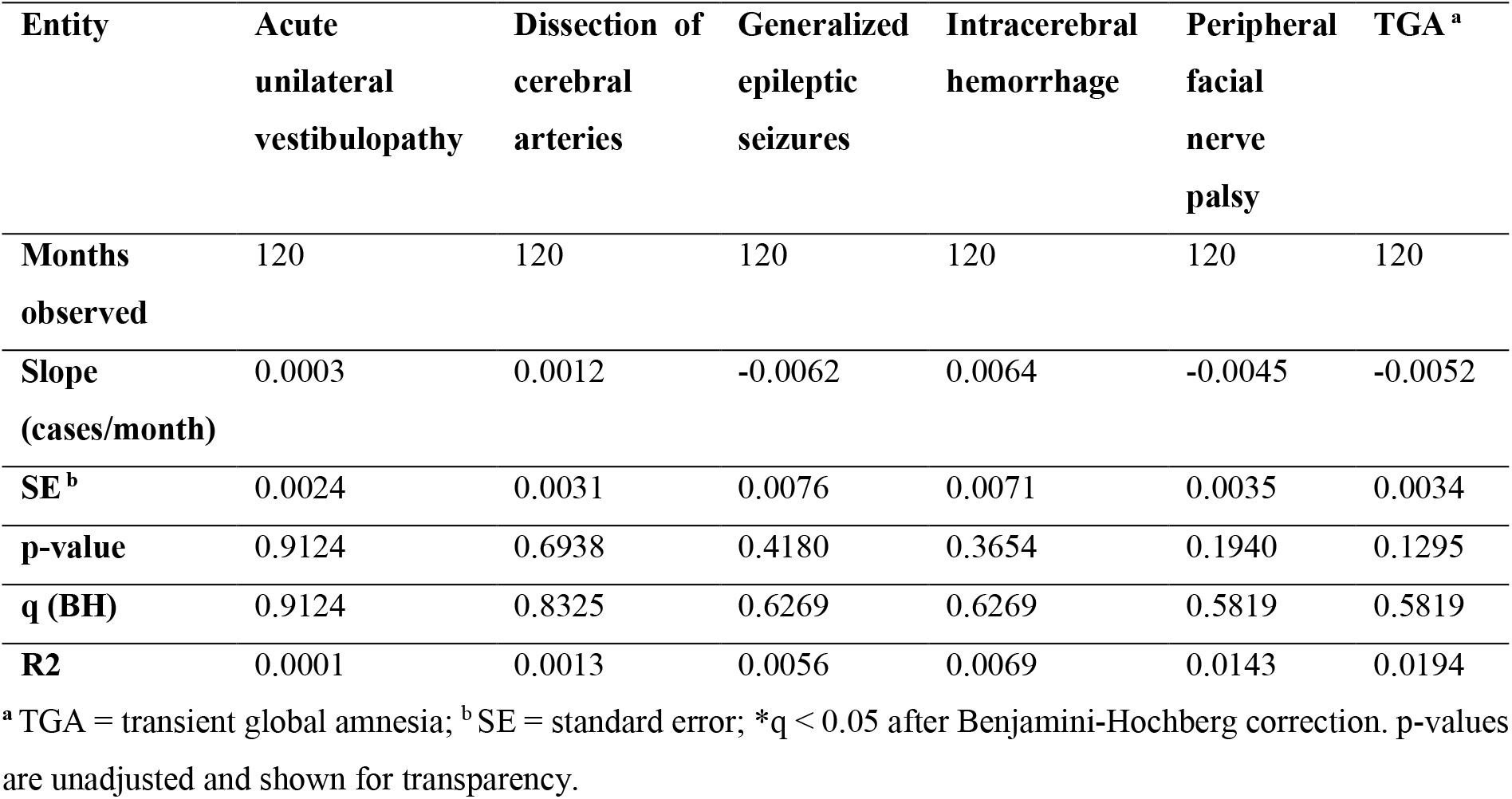
Regression analysis of the monthly distribution of admissions by diagnostic group.

### Monthly and Seasonal Distribution

Admissions for generalized epileptic seizures differed statistically significant from uniform monthly distribution (χ² = 33.7, df = 11, p < 0.001, and q = 0.0024), with a marked peak in January and a dip in May (Table 3, Figure 2), which corresponds to the observed winter predominance in absolute case counts (Table 4). In addition to that, an annual seasonal rhythm was identified in generalized epileptic seizures by Cosinor analysis (F = 3.37, p = 0.012; amplitude 1.09 cases/month, acrophase in November; Table 5) which did not hold up to correction for multiple comparisons (q = 0.0723).

**Figure 2.**
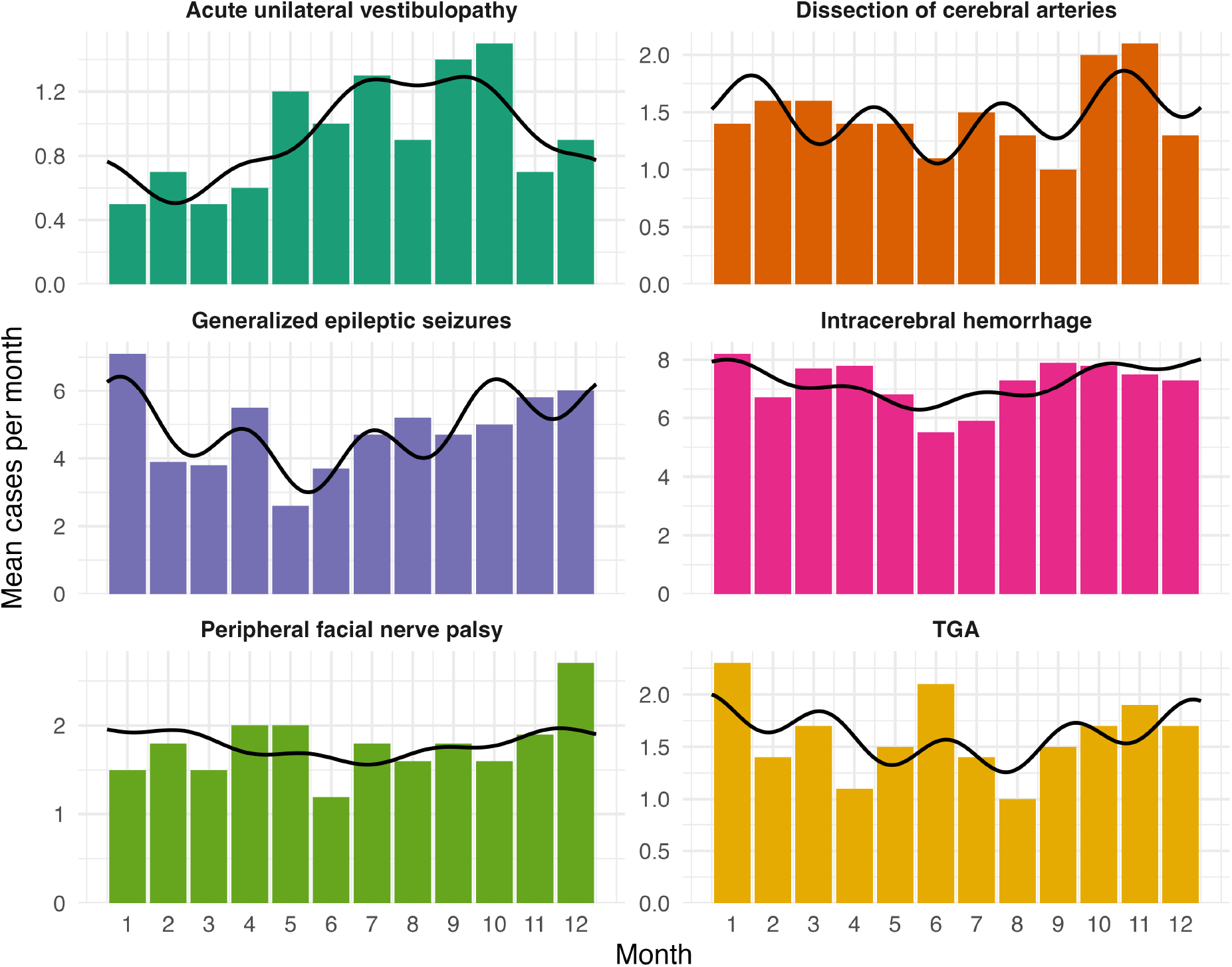
Monthly distribution of case counts and the seasonal fit derived from cosinor analysis for each diagnostic group. Generalized epileptic seizures and acute unilateral vestibulopathy showed an annual rhythmicity that did not survive correction for multiple comparisons; no significant seasonality was detected for the remaining four groups (see Tables 3 and 4), the fitted curves are shown descriptively.

**Table 3.** Chi-squared goodness-of-fit test for uniform monthly distribution of case counts by diagnostic group.

| Entity | Acute unilateral vestibulopathy | Dissection of cerebral arteries | Generalized epileptic seizures | Intracerebral hemorrhage | Peripheral facial nerve palsy | TGA <sup>a</sup> |
| --- | --- | --- | --- | --- | --- | --- |
| **Chi-square** | 14.43 | 7.75 | 33.70 | 10.50 | 8.50 | 9.76 |
| **df <sup>b</sup>** | 11 | 11 | 11 | 11 | 11 | 11 |
| **p-value** | 0.2102 | 0.7359 | 0.0004 | 0.4860 | 0.6675 | 0.5524 |
| **q (BH) <sup>c</sup>** | 0.6305 | 0.7359 | 0.0024* | 0.7359 | 0.7359 | 0.7359 |

<sup>a</sup> TGA = transient global amnesia; <sup>b</sup> df = degrees of freedom; <sup>c</sup> q (BH) = p-value adjusted for multiple comparisons across the six diagnostic groups using the Benjamini-Hochberg false discovery rate procedure. \*q < 0.05 after Benjamini-Hochberg correction. p-values are unadjusted and shown for transparency.

**Table 4.** Seasonal distribution of case counts by diagnostic group.

| Entity | Acute unilateral vestibulopathy | Dissection of cerebral arteries | Generalized epileptic seizures | ICH <sup>a</sup> | Peripheral facial nerve palsy | TGA <sup>b</sup> |
| --- | --- | --- | --- | --- | --- | --- |
| **Winter** | 21 | 43 | 170 | 222 | 60 | 54 |
| **Spring** | 23 | 44 | 119 | 223 | 55 | 43 |
| **Summer** | 32 | 39 | 136 | 187 | 46 | 45 |
| **Autumn** | 36 | 51 | 155 | 232 | 53 | 51 |
| **Total** | 112 | 177 | 580 | 864 | 214 | 193 |

<sup>a</sup> ICH = intracerebral hemorrhage; <sup>b</sup> TGA = transient global amnesia

**Table 5.** Cosinor analysis of seasonal rhythm by disease entity.

| Entity | Acute<br>unilateral<br>vestibulopathy | Dissection<br>of cerebral<br>arteries | Generalized<br>epileptic<br>seizures | Intracerebral<br>hemorrhage | Peripheral<br>facial nerve<br>palsy | TGA <sup>a</sup> |
| --- | --- | --- | --- | --- | --- | --- |
| **F-statistic** | 2.64 | 0.94 | 3.37 | 1.12 | 0.28 | 0.62 |
| **df <sup>b</sup>** | 4.114 | 4.114 | 4.114 | 4.114 | 4.114 | 4.114 |
| **p-value** | 0.0376 | 0.4446 | 0.0121 | 0.3529 | 0.8888 | 0.6484 |
| **q (BH) <sup>c</sup>** | 0.1128 | 0.6669 | 0.0723 | 0.6669 | 0.8888 | 0.7781 |
| **Amplitude,**<br>**annual**<br>**(cases/month)** | 0.36 | - | 1.09 | - | - | - |
| **Peak month**<br>**(annual cycle)** | Aug | - | Nov | - | - | - |
| **Amplitude,**<br>**quarterly**<br>**(cases/month)** | 0.06 | - | 0.75 | - | - | - |

<sup>a</sup> TGA= transient global amnesia; <sup>b</sup> df = degrees of freedom; <sup>c</sup> q (BH) = p-value adjusted for multiple comparisons across the six diagnostic groups using the Benjamini-Hochberg false discovery rate procedure. \*q < 0.05 after Benjamini-Hochberg correction. p-values are unadjusted and shown for transparency. Amplitude and acrophase are not reported where the cosinor model was not statistically significant within the p-values.

Acute unilateral vestibulopathy likewise showed an annual rhythm with an acrophase in August (F = 2.64, p = 0.038) that did not survive correction for multiple comparisons (q = 0.113; Table 5); no deviation from monthly distribution was detectable. (χ² = 14.43, p = 0.2102; Table 3). For the other four diagnostic groups (cerebral artery dissection, non-traumatic intracerebral hemorrhage, peripheral facial nerve palsy, or transient global amnesia) no monthly or seasonal rhythmicity was detected (all p > 0.05; Table 5, Figure 2).

### Temporal Clustering and Monte Carlo simulations

The mean inter-event interval differed greatly between diagnostic groups, ranging from 4.21 days in non-traumatic intracerebral hemorrhage to 31.77 days in acute unilateral vestibulopathy (Table 6, Figure 3a-b). The coefficient of variation of inter-event intervals exceeded 1 for generalized epileptic seizures (1.22), peripheral facial nerve palsy (1.23) and transient global amnesia (1.03), consistent with over-dispersion relative to a homogeneous Poisson process. Non-traumatic intracerebral hemorrhage was indistinguishable from Poisson (1.00), whereas acute unilateral vestibulopathy (0.87) and cerebral artery dissection (0.95) showed values below 1, indicating a more regular than random spacing of events.

**Figure 3.**
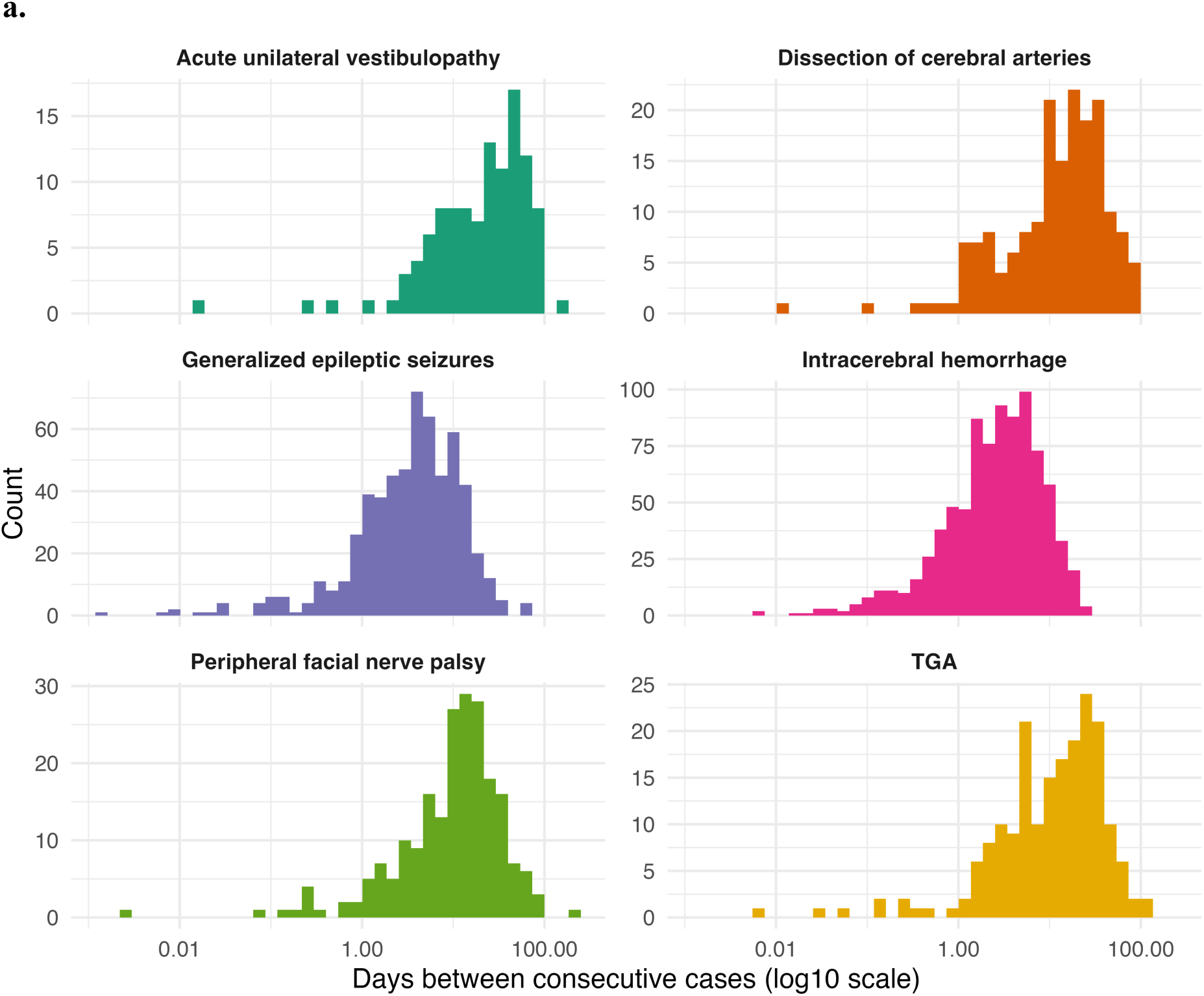
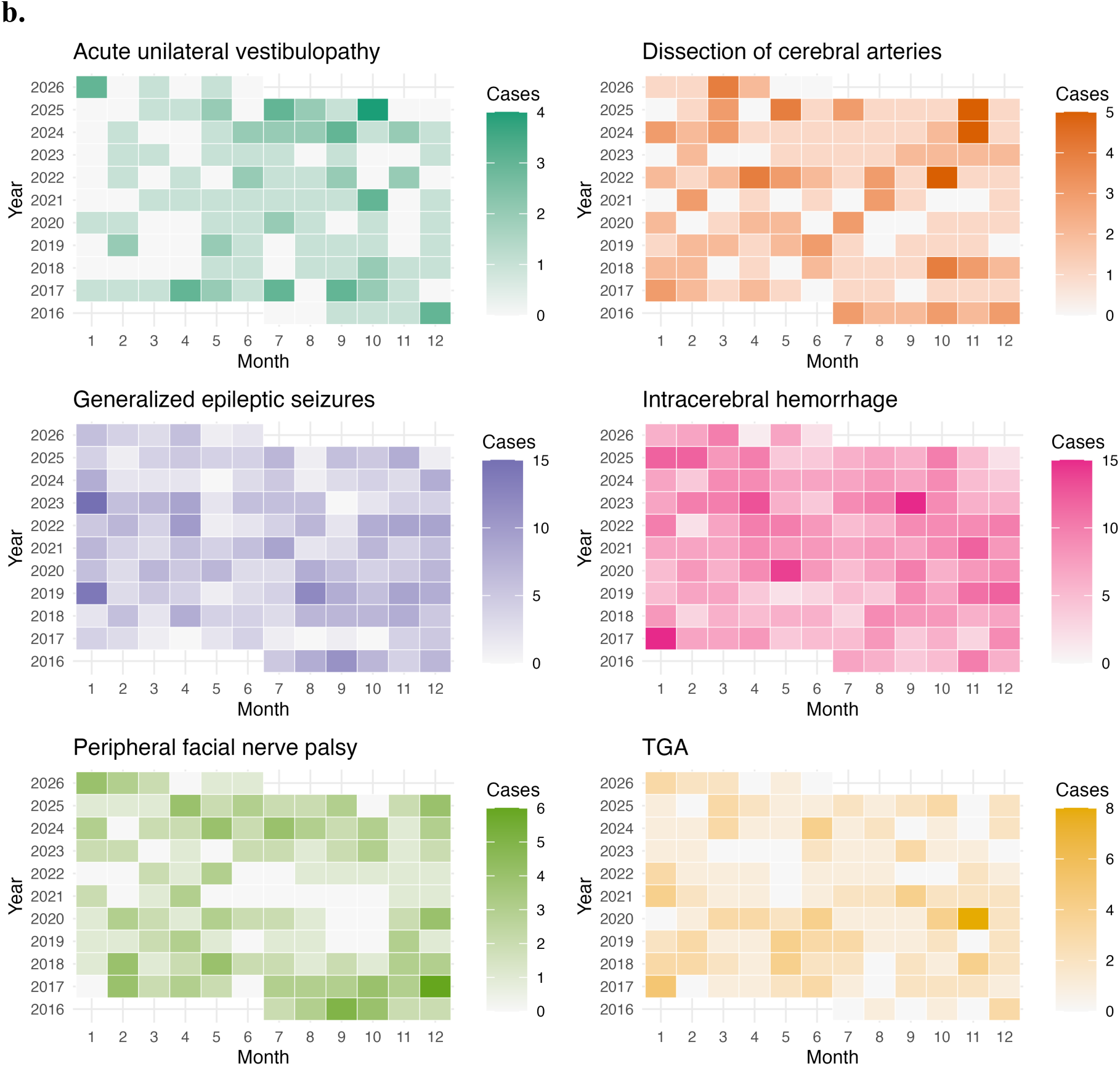
(a) Distribution of inter-event intervals (log scale) by disease entity, illustrating the shape of the time-gap distribution between consecutive events per diagnostic group. (b) Heatmap of monthly case counts by calendar year across all six diagnostic groups over the observed 10-year period, visualizing the temporal distribution of cases across the observation period.

**Table 6.** Inter-event interval statistics and coefficient of variation by diagnostic group.

| Entity | Acute<br>unilateral<br>vestibulopathy | Dissection of<br>cerebral<br>arteries | Generalized<br>epileptic<br>seizures | Intracerebral<br>hemorrhage | Peripheral<br>facial nerve<br>palsy | TGA<sup>a</sup> |
| --- | --- | --- | --- | --- | --- | --- |
| **Mean IEI<sup>b</sup>**<br>**(days)** | 31.77 | 20.34 | 6.26 | 4.21 | 17.01 | 18.55 |
| **SD<sup>c</sup> (days)** | 27.71 | 19.28 | 7.65 | 4.23 | 20.92 | 19.16 |
| **Coefficient of**<br>**variation**<br>**(CV)** | 0.87 | 0.95 | 1.22 | 1.00 | 1.23 | 1.03 |

<sup>a</sup> TGA = transient global amnesia; <sup>b</sup> IEI = inter-event interval; <sup>c</sup> SD = standard deviation

After correction for multiple comparisons, the Monte Carlo simulations using n = 999 permutations showed significant temporal clustering for three of the six diagnostic groups (Figures 4a-f): generalized epileptic seizures, peripheral facial nerve palsy, and transient global amnesia (Table 7). The complete results for all six diagnostic groups and six time windows are in given in Supplementary Table 1.

**Figure 4.**
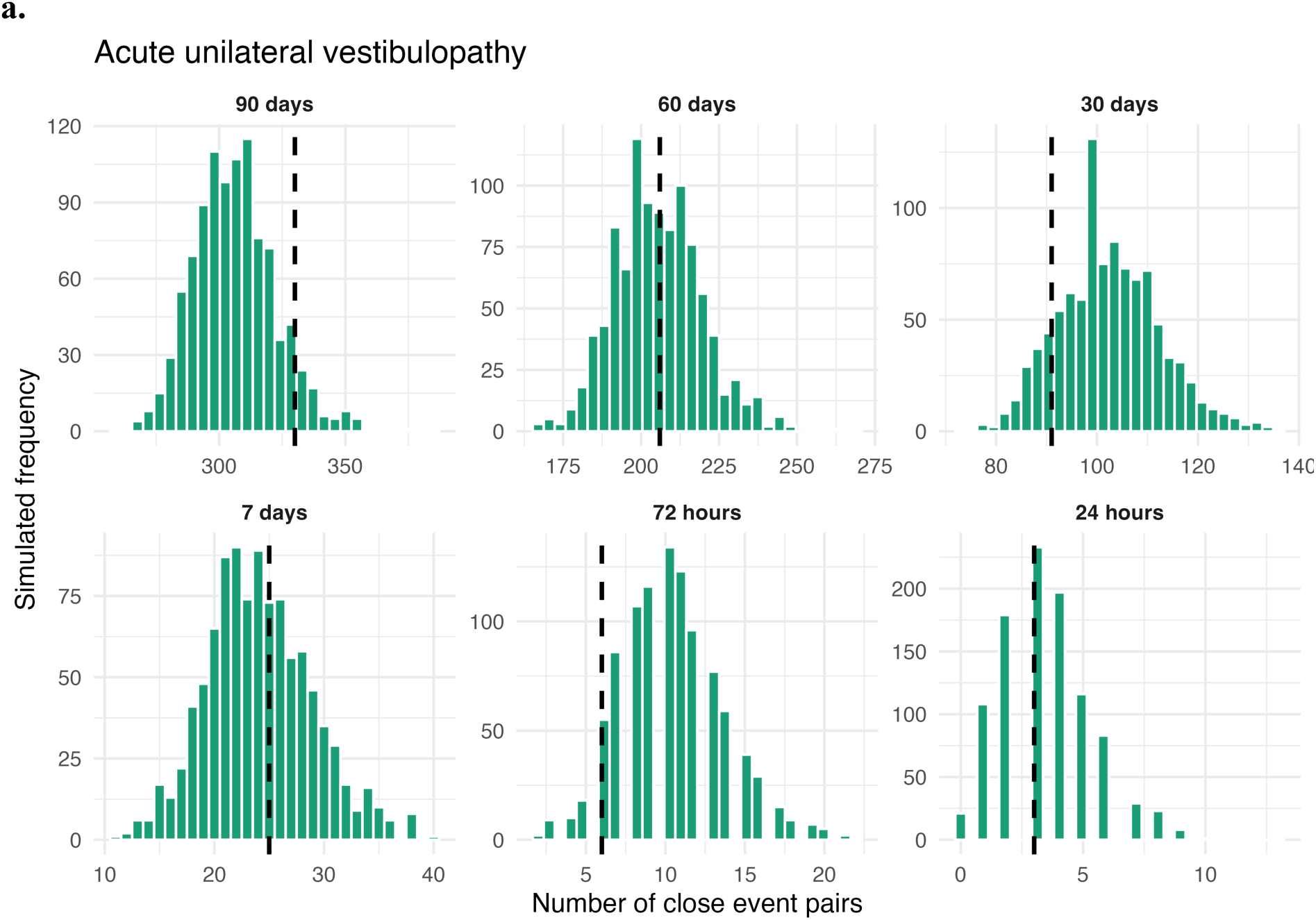
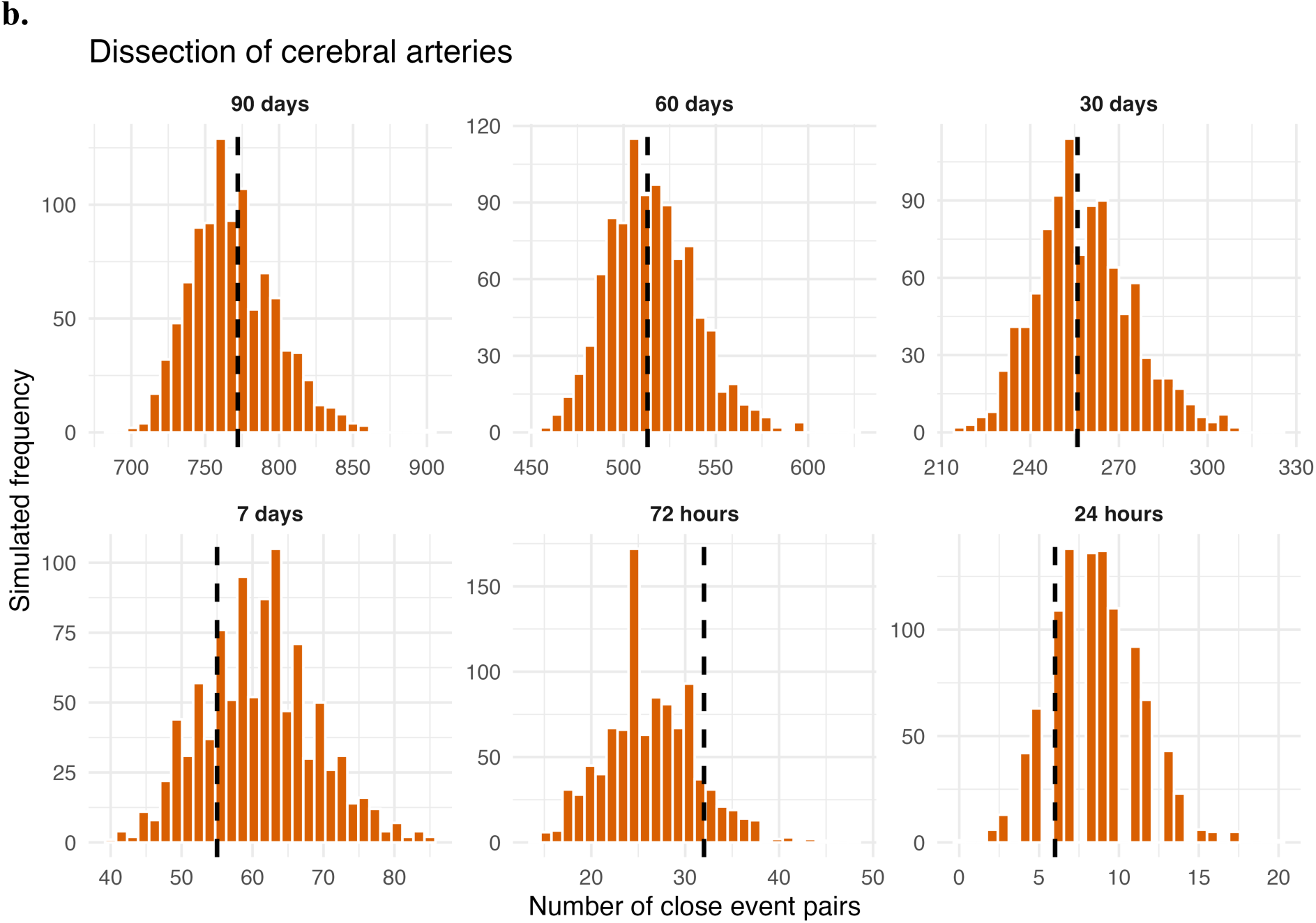
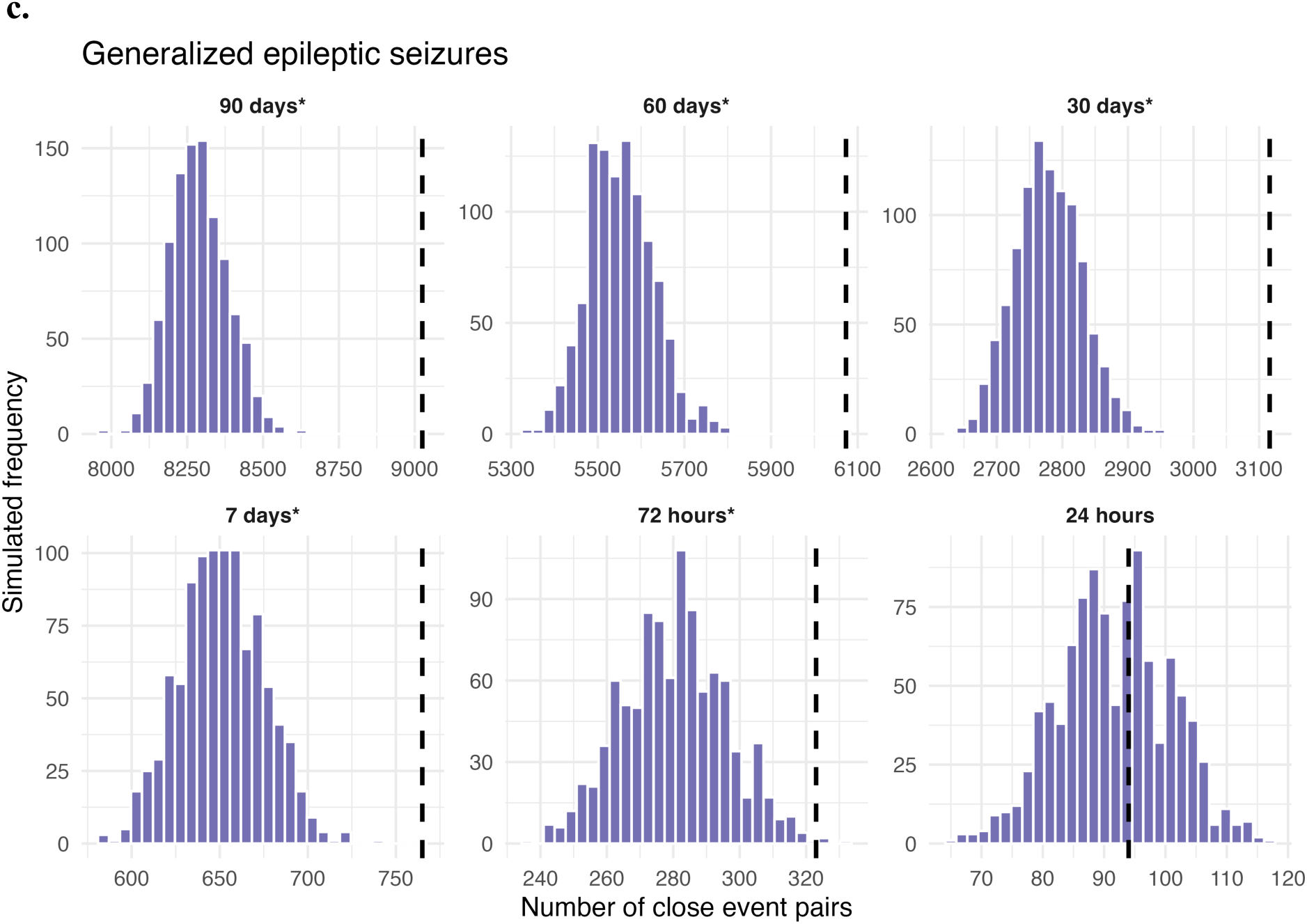
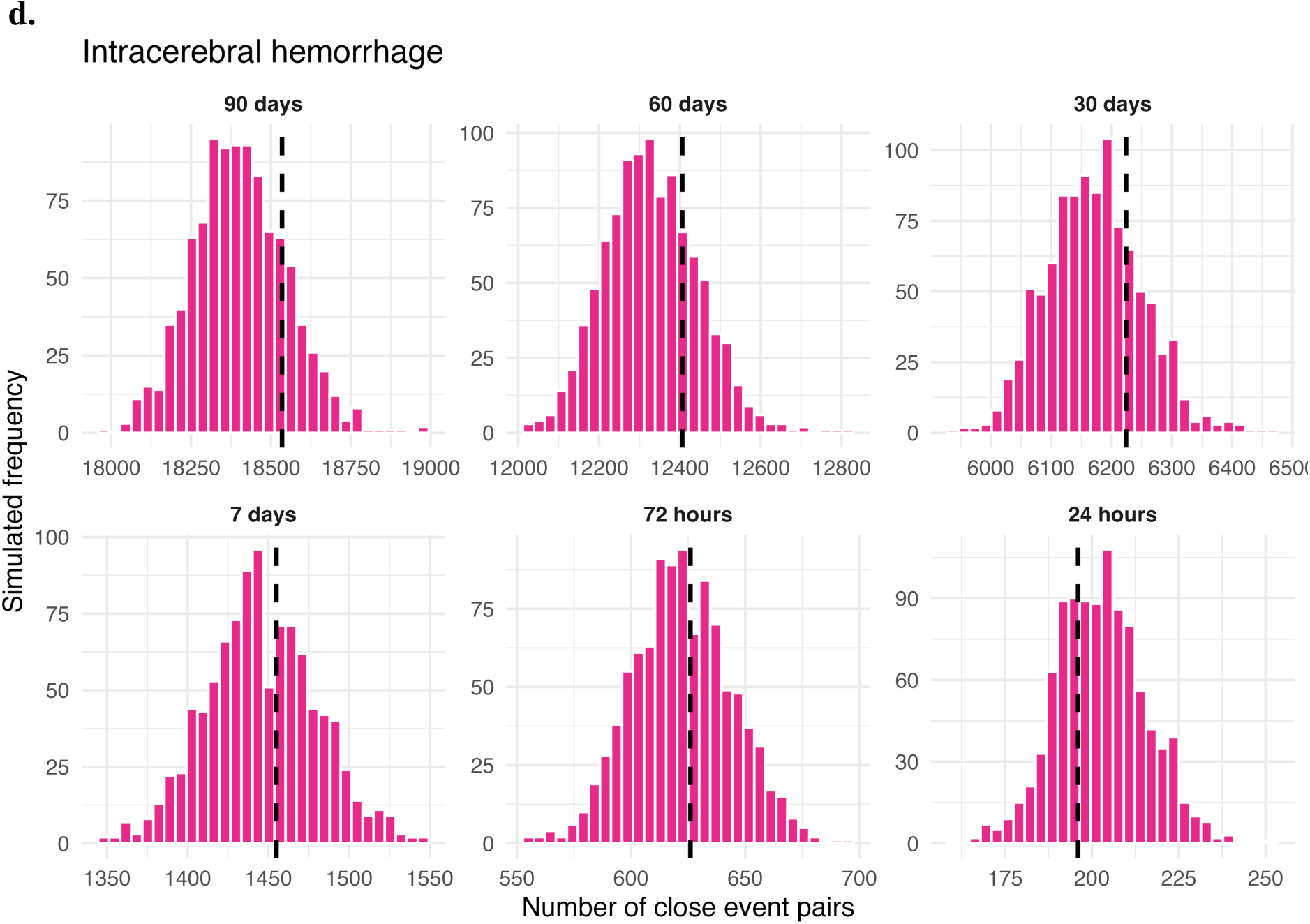
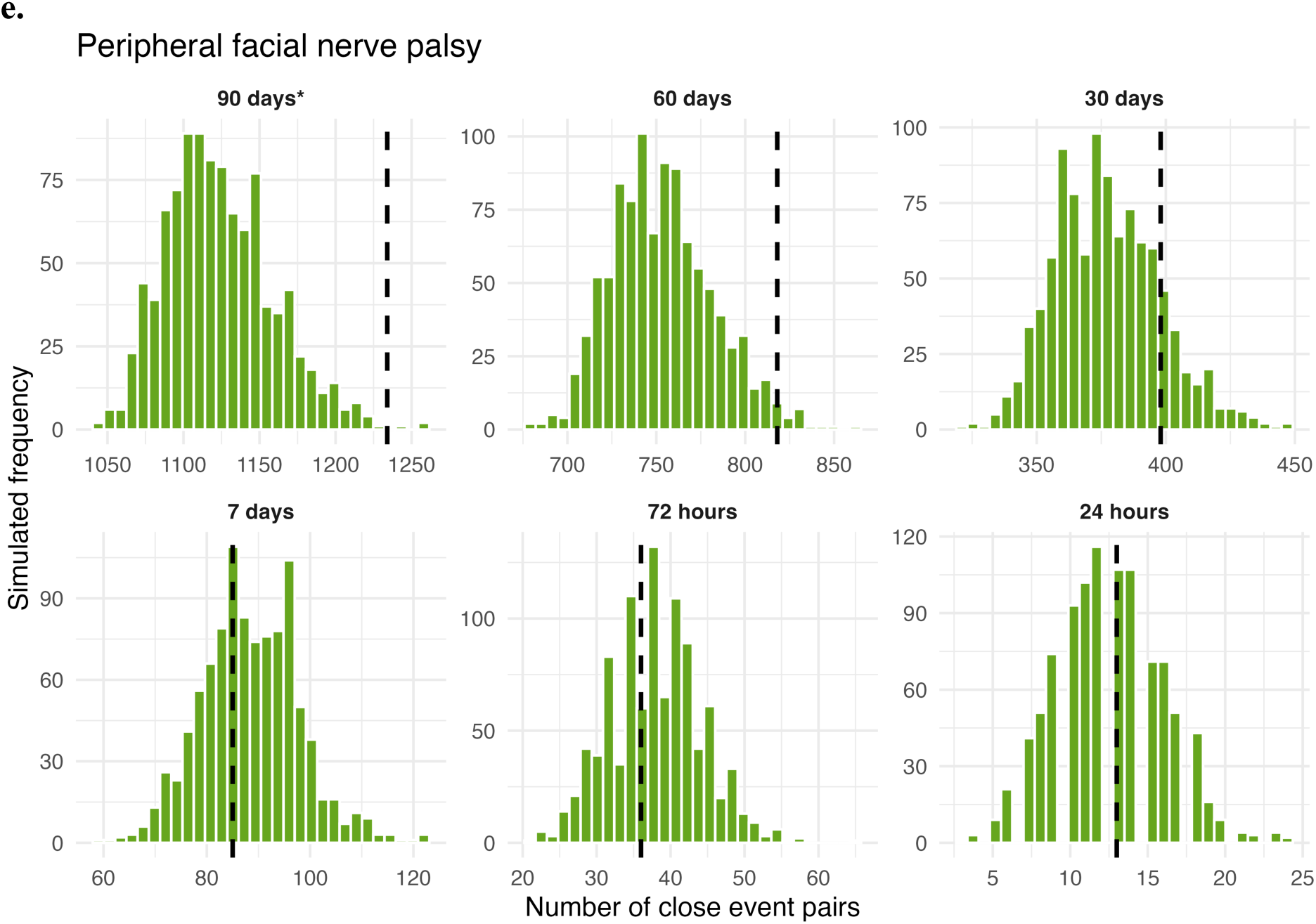
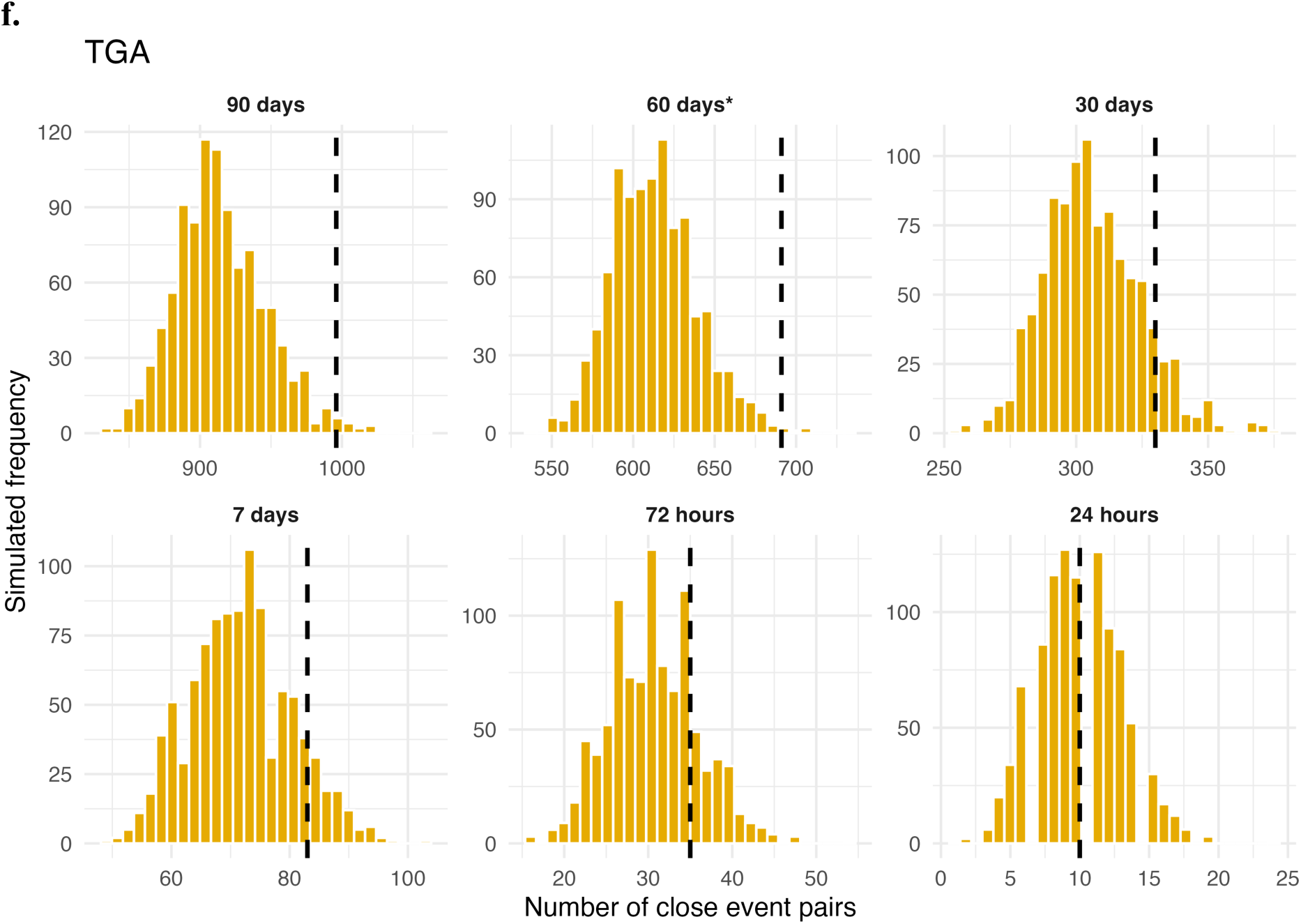
**(a–f).** Monte Carlo permutation null distributions (999 permutations) versus observed close-event pair counts across time windows (24 hours - 90 days), shown separately for each of the six diagnostic groups: (a) acute unilateral vestibulopathy, (b) dissection of cerebral arteries, (c) generalized epileptic seizures, (d) intracerebral hemorrhage, (e) peripheral facial nerve palsy, (f) TGA.

**Table 7.** Results of Monte Carlo permutation testing for temporal clustering.

| Entity | Time window | Observed close pairs | Expected (MC mean) | MC SD<sup>a</sup> | Standardized score | p-value (Monte Carlo) | q (BH, 36 tests)<sup>c</sup> |
| --- | --- | --- | --- | --- | --- | --- | --- |
| **Generalized epileptic seizures** | 90 days | 9025 | 8288.9 | 94.4 | 7.80 | 0.001 | 0.009* |
| **Generalized epileptic seizures** | 60 days | 6073 | 5553.7 | 75.4 | 6.88 | 0.001 | 0.009* |
| **Generalized epileptic seizures** | 30 days | 3116 | 2778.9 | 51.9 | 6.50 | 0.001 | 0.009* |
| **Generalized epileptic seizures** | 7 days | 765 | 651.1 | 24.0 | 4.75 | 0.001 | 0.009* |
| **Generalized epileptic seizures** | 72 hours | 323 | 280.3 | 15.6 | 2.74 | 0.005 | 0.030* |
| **Peripheral facial nerve palsy** | 90 days | 1234 | 1123.8 | 35.1 | 3.14 | 0.004 | 0.029* |
| **Peripheral facial nerve palsy** | 60 days | 818 | 753.4 | 28.5 | 2.27 | 0.022 | 0.088 |
| **TGA<sup>b</sup>** | 90 days | 996 | 914.9 | 32.0 | 2.54 | 0.019 | 0.085 |
| **TGA<sup>b</sup>** | 60 days | 691 | 613.2 | 26.7 | 2.92 | 0.008 | 0.041* |

<sup>a</sup>MC = Monte Carlo permutation test (999 permutations). Expected values and SD derived from permutation null distribution. Standardized score = (observed - expected) / Monte Carlo SD; <sup>b</sup>TGA = transient global amnesia; <sup>c</sup>q = p-value adjusted for multiple comparisons across all 36 tests (six diagnostic groups × six time windows) using the Benjamini-Hochberg false discovery rate procedure. Only time windows with p < 0.05 are shown, p-values are unadjusted and shown for transparency. \*q < 0.05 after Benjamini-Hochberg correction. Complete results for all time windows and diagnostic groups are provided in Supplementary Table S1.

Robust clustering across all time windows ranging from 72 hours to 90 days (all p < 0.01, q ≤ 0.03) was detected in the incidence of generalized epileptic seizures, surpassing the expected Monte Carlo values (MC mean) of observed close-event pairs consistently and substantially. Yet no statistically significant clustering effect was identified at the 24-hour time window (p = 0.425), suggesting a clustering in the incidence of generalized epileptic seizures on a weekly to monthly timescale rather than within individual days.

Peripheral facial nerve palsy showed a significant temporal clustering in the 90-day (p = 0.004, q = 0.029) time window, the 60-day window reached nominal significance, but did not survive multiple comparisons correction (p = 0.022, q = 0.088). Shorter time periods showed no temporal clustering (all p > 0.05), proposing a possible environmental or seasonal trigger consistent with medium-term temporal clustering.

The data for admission of transient global amnesia, unlike the peripheral facial nerve palsy, showed in the Monte Carlo permutation testing a significant clustering for the 60-day (p = 0.008, q = 0.041) time window, the 90-day time frame did not withstand the multiple comparison correction (p = 0.008, q = 0.041). No excess temporal clustering was detected at shorter time windows (all p >0.05).

Across any time windows no observation of significant temporal clustering was made for acute unilateral vestibulopathy, dissection of cerebral arteries, or non-traumatic intracerebral hemorrhage (all p > 0.05).

### Endogenous cluster dynamics (Hawkes process modelling)

Hawkes process modelling showed no statistically self-excitation for any of the diagnostic groups after correction for multiple comparisons. Generalized epileptic seizures showed the largest effect (likelihood ratio test statistic 6.582 with a p-value = 0.036, q = 0.216), the estimated excitation magnitude was α = 0.40589 with a decay rate of β = 32.29062/ day, corresponding to an excitation half-life of 31 minutes. Fewer than 8 of the 580 admissions over the analyzed 10-year period were attributable to a self-excitation point process, the temporal clustering identified by Monte Carlo permutation testing is thus not adequately captured by a self-excitation point process with a single exponential root (Table 8, Figure 5).

**Figure 5.**
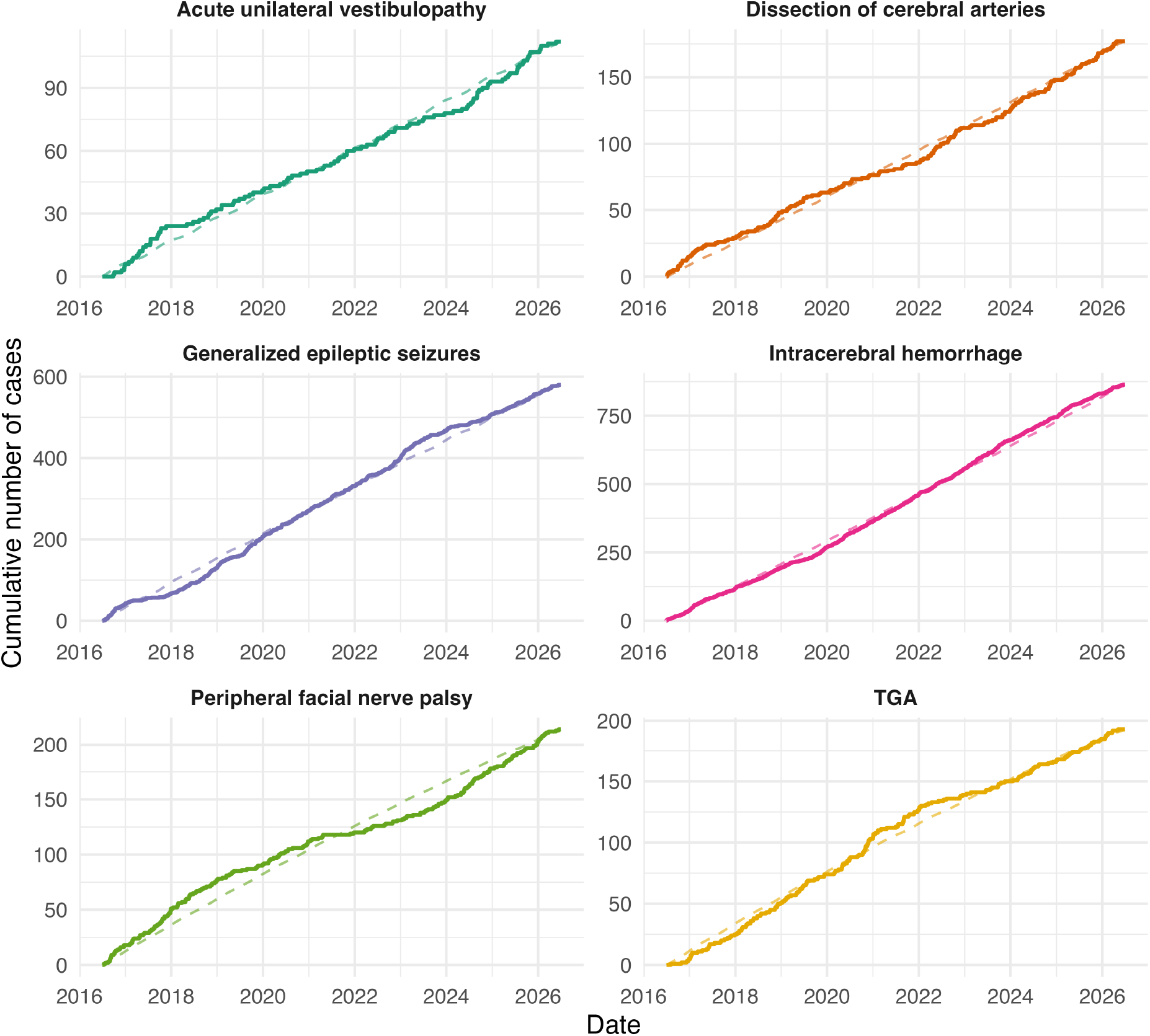
Cumulative number of cases over the observation period for each of the six diagnostic groups: acute unilateral vestibulopathy, dissection of cerebral arteries, generalized epileptic seizures, intracerebral hemorrhage, peripheral facial nerve palsy, and TGA. Solid line: observed cumulative case count; dashed line: expected cumulative count under the seasonality-adjusted background rate (incorporating the annual rhythm derived from cosinor analysis). Deviations of the observed from the expected trajectory hint at possible temporal clustering beyond what is attributable to seasonal variation alone; no diagnostic group showed statistically significant self-excitation after correction for multiple comparisons (see Table 8).

**Table 8.** Hawkes process parameter estimates and self-excitation test results by diagnostic group.

| Entity | Acute<br>unilateral<br>vestibulopathy | Dissection<br>of<br>cerebral<br>arteries | Generalized<br>epileptic<br>seizures | Intracerebral<br>hemorrhage | Peripheral<br>facial nerve<br>palsy | TGA |
| --- | --- | --- | --- | --- | --- | --- |
| **n** | 112 | 177 | 580 | 864 | 214 | 193 |
| **$\alpha$**<br>(excitation) | 0.54843 | 0.38358 | 0.40512 | 0.00010 | 0.06481 | 0.20197 |
| **$\beta$** (decay<br>rate/day) | 71.98209 | 89.04709 | 32.44643 | 0.01428 | 5.83807 | 10.48434 |
| **Branching**<br>**ratio** | 0.0076 | 0.0043 | 0.0125 | 0.0072 | 0.0111 | 0.0193 |
| **Excitation**<br>**half-life**<br>**(days)** | 0.0096 | 0.0078 | 0.0214 | 48.5298 | 0.1187 | 0.0661 |
| **LRT**<br>**statistic <sup>a</sup>** | 2.133 | 1.341 | 6.536 | 0.046 | 0.954 | 4.087 |
| **p**<br>(bootstrap<br>LRT) | 0.2390 | 0.3210 | 0.0240 | 0.6600 | 0.4040 | 0.0750 |
| **q (BH) <sup>b</sup>** | 0.4780 | 0.4815 | 0.1440 | 0.6600 | 0.4848 | 0.2250 |

<sup>a</sup> LRT = likelihood ratio test; <sup>b</sup> q = p-value adjusted for multiple comparisons across the six diagnostic groups using the Benjamini-Hochberg false discovery rate procedure.

No statistically self-excitation was detected for any of the remaining five diagnostic groups (p > 0.05; Table 8, Figure 5).

## Discussion

Generalized epileptic seizures were the only diagnostic group that showed a significant seasonal variation, significant deviation from a uniform monthly distribution, and significant temporal clustering. Within the admissions of peripheral facial palsy and transient global amnesia this study was able to detect medium-term temporal clustering without seasonality, whereas no significant temporal clustering or seasonality was observed for dissection of cerebral arteries, non-traumatic intracerebral hemorrhage, and acute unilateral vestibulopathy. Generalized epileptic seizures were the only diagnostic group that showed significant seasonal variation and temporal clustering.

These findings suggest that the incidence of acute neurological conditions is not uniformly distributed across time, and disease expression may be modulated by environmental, infectious, and potentially endogenous biological factors.

Whilst infectious triggers, particularly viral infection, may precipitate seizures, population-level seasonality data has been inconsistent^20^ and the relationship between generalized epileptic seizures and seasonality is complex and less well established than for vascular conditions. In the present study, a statistically significant seasonal pattern in seizure incidence was nonetheless observed. The peak monthly count fell in January, while the acrophase derived from cosinor analysis was obtained in November. This timing raises the possibility of an infectious trigger, given that colder months are typically associated with closer person-to-person contact and, in turn, more rapid transmission of viral illness. A further finding of the present study was the presence of temporal clustering at comparatively large time scales, ranging from 72 hours to 90 days, which was not accompanied by a significant self-excitation point process in Hawkes process modelling. A temporal clustering reflecting synchronized generalized epileptic seizures across different patients points toward shared, population-level triggers, e.g. the above-mentioned viral infection, sleep disruption or altered alcohol consumption could conceivably act on many patients simultaneously if driven by shared seasonal exposures (e.g. holiday-related alcohol intake, seasonal viral epidemics).

For acute unilateral vestibulopathy, there has been only little systematic investigation of potential seasonal variation in the incidence, despite a viral or post-viral inflammatory mechanism being assumed as a possible etiology. The present study cosinor analysis fitted an annual rhythm with the acrophase falling in August, which, however, did not survive the correction for multiple comparisons, and, in addition to that, the monthly distribution did not differ from uniform. Given the presumed viral or post-viral etiology of the condition, the absence of a winter or spring predominance is not analogous to other post-infectious neurological syndromes; for exclusion of a moderate seasonal effect, a larger cohort would be needed than in the present study (n = 112).

Peripheral facial nerve palsy has previously been reported to occur more frequently during colder months, potentially linked to viral reactivation under conditions of cold exposure and immune modulation^17–19^. In contrast, our study found no deviation from uniform seasonal distribution. This is compatible with previous findings that have not been entirely consistent across geographic regions and climate zones, as illustrated by a recent systematic review that portrayed substantial heterogeneity in reported seasonal patterns^18^. Nevertheless, a significant temporal clustering was present within the 90-day time scale, suggesting that occurrence is not randomly distributed over time despite the absence of an annual pattern. Such clustering may be compatible with reactivation mechanisms triggered by non-calendar or non-seasonal factors, such as stress or episodic cold exposure.

The absence of statistically significant seasonality in transient global amnesia is consistent with the largest and best-powered cohort published to date, a two-center study of 665 patients from Mannheim, Germany, and the Kansai district, Japan, which also found no significant variation by month or season ^31^. Several smaller, single-center studies reported seasonal peaks ^32–34^, yet there was no consistency in the reported peak months across studies, which samples the heterogenous distribution consistent with the absence of seasonality reported here. The absence of seasonality argues against a population-level, climate-driven mechanism and is more in line with the episodic, individual trigger-dependent presentation, which is classically described for TGA; the pathophysiology hypothesis of stress-related cortical spreading depression mechanisms and transient hippocampal ischemia precipitated by venous congestion through Valsalva-associated retrograde flow in the internal jugular veins ^35^, and itself being precipitated by physical and/or emotional stress, is congruent with a non-seasonal occurrence and could cluster in time if arising from shared but irregularly timed population-level events.

A predilection for the colder months in the incidence of non-traumatic intracerebral hemorrhage has been reported in multiple prior studies ^1, 2, 4, 5, 8^, possibly mediated by cold-induced blood pressure elevation ^11^. In the present study, however, non-traumatic intracerebral hemorrhage showed neither significant seasonality nor temporal clustering. This may be more consistent with pathophysiological processes operating predominantly at the individual patient level rather than with a population-level change. This fits previously identified possible precipitants for primary intracerebral bleeding, including Valsalva maneuvers, exertion and activity, and emotional stress ^36^.

Contrasting to several previous reports of a winter predominance in cervical artery dissection^24–27^, the study present could not identify a statistically significant seasonality for dissections of the cervical and intracranial arteries. No significant annual rhythm was detected for cerebral artery dissection, and the acrophase estimated from a non-significant model is not interpretable. The absence of the cold-season predominance reported in cervical artery dissection cohorts may reflect the composition of the present group, which combined extracranial and intracranial dissections. A significant seasonality in one subgroup, if the two subtypes differ in their underlying triggers, could be masked in a joint analysis. Studies with sufficient power to stratify by dissection site will be needed to determine whether the seasonality reported for cervical artery dissection specifically also holds when cervical and intracranial cases are considered separately.

Taken together, the heterogeneous temporal patterns observed across the six diagnostic groups suggest that seasonality and temporal clustering represent distinct dimensions of temporal disease occurrence. Generalized epileptic seizures uniquely combined both, temporal clustering and seasonality, whereas peripheral facial nerve palsy and TGA showed episodic clustering without an annual rhythm; no temporal clustering, seasonality or self-excitation process was present in acute unilateral vestibulopathy, non-traumatic intracerebral hemorrhage and dissection of cerebral arteries.

These findings indicate that seasonality analysis alone may be insufficient to capture the temporal architecture of acute neurological events. Combining analyses of seasonal variation, temporal clustering, and self-excitation may provide additional insight into whether observed temporal patterns are compatible with recurring environmental exposures, irregular population-level triggers, or processes predominantly operating at the individual patient level.

The observed patterns also provide a quantitative perspective on the clinical impression that certain acute neurological diagnoses appear to occur in “theme days” or “theme weekends”. Importantly, however, the present findings suggest that such temporal patterning is not a universal feature of acute neurological disease but appears to be confined to specific conditions.

Several methodological aspects of this study should be considered together when interpreting the findings. The retrospective, single-center design necessarily limits the generalizability of the results; at the same time, studying a 10-year period within a single tertiary neurological center provides a large and internally consistent dataset with relatively stable diagnostic and administrative structures. The simultaneous analysis of six clinically distinct acute neurological conditions using an identical analytical framework further enables direct within-study comparisons of temporal patterns across disease entities.

The use of the timestamp of hospital admission as a proxy for symptom onset may have introduced temporal misclassification because delays between symptom onset and admission can occur. However, such delays would be unlikely to vary systematically by season or across the temporal clustering windows examined and would therefore be expected predominantly to reduce rather than artificially generate temporal signals.

Several relevant patient- and population-level variables were unavailable. Patient-level risk factors such as smoking, chronic kidney disease, and heart failure were inconsistently recorded and could therefore not be adjusted for. Population-level infection surveillance data, which would be required to directly test several of the infectious-trigger hypotheses discussed above, were also unavailable. Disease-specific severity measures were likewise unavailable, precluding subgroup analyses of whether disease severity or prognosis varies with onset timing.

Two more limitations should be noted: Firstly, the study period included the COVID-19 pandemic time window, during which admission volumes for acute neurological conditions changed substantially and non-linearly, secondly, the permutation null distribution preserves the integrity of temporal distribution of admissions but does not account for year-to-year changes in admissions within individual diagnostic groups. Temporal clustering present only at larger time frame may thus partly reflect slow variation in case numbers rather than short-term clustering; the findings for peripheral facial nerve palsy and transient global amnesia should be deemed as hypothesis-generating.

A methodological strength of the present study is the multi-layered analytical framework used to address these different aspects of temporal structure. A chi-squared goodness-of-fit test was used for initial screening of deviations from uniform monthly incidence, while cosinor analysis provided quantitative estimates of rhythmic seasonal patterns, including amplitude and acrophase. Monte Carlo permutation analysis was used to assess short-term temporal clustering by comparing observed inter-event intervals with a synthetic null distribution. Finally, Hawkes self-exciting point process modeling was applied to characterize the dynamics of identified clusters and estimate background event rates, excitation magnitude, and decay. Long-term seasonal fluctuations derived from cosinor analysis were integrated into the background rate component of the Hawkes model to separate short-term excitation effects from seasonal confounding, the permutation analysis tests against the pooled temporal structure of all admissions and is not adjusted for group-specific seasonality.

To our knowledge, this combined analytical approach has not previously been applied to the simultaneous investigation of multiple acute neurological conditions and therefore represents a methodological contribution of the present study.

## Conclusions

The present study demonstrates disease-specific temporal patterning across six acute neurological conditions. After correction for multiple comparisons, generalized epileptic seizures was the only diagnostic group that uniquely combined significant seasonality and temporal clustering, whereas peripheral facial nerve palsy and transient global amnesia showed medium-term clustering in the absence of annual seasonality. Acute unilateral vestibulopathy, cerebral artery dissection and non-traumatic intracerebral hemorrhage showed neither seasonality nor clustering, no diagnostic group showed significant self-excitation.

The anecdotal impression of diagnostic “theme shifts” among on-call neurologists therefore appears to have a measurable basis, although clustering is confined to specific conditions. These findings are hypothesis-generating, and the mechanisms underlying these disease-specific temporal patterns remain to be determined in larger, multicenter studies incorporating patient-level and population-level exposures.

## Data Availability

All data produced in the present study are available upon reasonable request to the authors.

---

**Supplementary Table 1:** Results of Monte Carlo permutation testing for temporal clustering distinguished by diagnostic group and time window.

| Entity | Time window | Observed close pairs | Expected (MC mean) | MC SD | Standardized score | p-value (Monte Carlo) | q (BH, 36 tests) <sup>e</sup> |
| --- | --- | --- | --- | --- | --- | --- | --- |
| AUV <sup>a</sup> | 90 days | 330 | 306.4 | 16.8 | 1.41 | 0.083 | 0.299 |
| AUV <sup>a</sup> | 60 days | 206 | 205.4 | 14.4 | 0.04 | 0.483 | 0.694 |
| AUV <sup>a</sup> | 30 days | 91 | 102.4 | 10.0 | -1.14 | 0.886 | 0.911 |
| AUV <sup>a</sup> | 7 days | 25 | 24.1 | 4.8 | 0.18 | 0.439 | 0.687 |
| AUV <sup>a</sup> | 72 hours | 6 | 10.4 | 3.2 | -1.38 | 0.961 | 0.961 |
| AUV <sup>a</sup> | 24 hours | 3 | 3.5 | 1.9 | -0.28 | 0.692 | 0.783 |
| Dissection of cerebral arteries | 90 days | 772 | 769.1 | 29.3 | 0.10 | 0.426 | 0.687 |
| Dissection of cerebral arteries | 60 days | 513 | 515.2 | 24.1 | -0.09 | 0.508 | 0.694 |
| Dissection of cerebral arteries | 30 days | 256 | 258.6 | 16.6 | -0.15 | 0.536 | 0.694 |
| Dissection of cerebral arteries | 7 days | 55 | 60.9 | 7.9 | -0.75 | 0.783 | 0.854 |
| Dissection of cerebral arteries | 72 hours | 32 | 26.3 | 5.0 | 1.14 | 0.148 | 0.410 |
| Dissection of cerebral arteries | 24 hours | 6 | 8.6 | 2.8 | -0.93 | 0.875 | 0.911 |
| Generalized Epileptic seizures | 90 days | 9025 | 8288.9 | 94.4 | 7.80 | 0.001 | 0.009* |
| Generalized Epileptic seizures | 60 days | 6073 | 5553.7 | 75.4 | 6.88 | 0.001 | 0.009* |
| Generalized Epileptic seizures | 30 days | 3116 | 2778.9 | 51.9 | 6.50 | 0.001 | 0.009* |
| Generalized Epileptic seizures | 7 days | 765 | 651.1 | 24.0 | 4.75 | 0.001 | 0.009* |
| Generalized Epileptic seizures | 72 hours | 323 | 280.3 | 15.6 | 2.74 | 0.005 | 0.030* |
| Generalized Epileptic seizures | 24 hours | 94 | 91.7 | 8.9 | 0.26 | 0.425 | 0.687 |
| ICH <sup>b</sup> | 90 days | 18535 | 18402.7 | 146.6 | 0.90 | 0.189 | 0.454 |
| ICH <sup>b</sup> | 60 days | 12406 | 12330.3 | 116.0 | 0.65 | 0.252 | 0.510 |
| ICH <sup>b</sup> | 30 days | 6224 | 6171.5 | 78.9 | 0.67 | 0.245 | 0.510 |
| ICH <sup>b</sup> | 7 days | 1455 | 1445.8 | 34.0 | 0.27 | 0.392 | 0.687 |
| ICH <sup>b</sup> | 72 hours | 626 | 622.5 | 21.9 | 0.16 | 0.436 | 0.687 |
| ICH <sup>b</sup> | 24 hours | 196 | 202.7 | 13.0 | -0.51 | 0.696 | 0.783 |
| **Peripheral facial nerve palsy** | 90 days | 1234 | 1123.8 | 35.1 | 3.14 | 0.004 | 0.029* |
| **Peripheral facial nerve palsy** | 60 days | 818 | 753.4 | 28.5 | 2.27 | 0.022 | 0.088 |
| **Peripheral facial nerve palsy** | 30 days | 398 | 377.1 | 20.4 | 1.02 | 0.163 | 0.419 |
| **Peripheral facial nerve palsy** | 7 days | 85 | 88.5 | 9.5 | -0.36 | 0.652 | 0.782 |
| **Peripheral facial nerve palsy** | 72 hours | 36 | 37.9 | 6.3 | -0.31 | 0.647 | 0.782 |
| **Peripheral facial nerve palsy** | 24 hours | 13 | 12.5 | 3.5 | 0.14 | 0.489 | 0.694 |
| **TGA<sup>c</sup>** | 90 days | 996 | 914.9 | 32.0 | 2.54 | 0.019 | 0.085 |
| **TGA<sup>c</sup>** | 60 days | 691 | 613.2 | 26.7 | 2.92 | 0.008 | 0.041* |
| **TGA<sup>c</sup>** | 30 days | 330 | 306.3 | 18.3 | 1.29 | 0.115 | 0.357 |
| **TGA<sup>c</sup>** | 7 days | 83 | 71.7 | 8.7 | 1.30 | 0.119 | 0.357 |
| **TGA<sup>c</sup>** | 72 hours | 35 | 30.8 | 5.4 | 0.79 | 0.255 | 0.510 |
| **TGA<sup>c</sup>** | 24 hours | 10 | 10.0 | 3.1 | 0.02 | 0.540 | 0.694 |

<sup>a</sup> AUV = acute unilateral vestibulopathy; <sup>b</sup> ICH = intracerebral hemorrhage; <sup>c</sup> TGA = transient global amnesia;
<sup>d</sup> MC = Monte Carlo permutation test (999 permutations). Expected values and SD derived from permutation null distribution. Standardized score = (observed – expected) / Monte Carlo SD; <sup>e</sup> q = p-value adjusted for multiple comparisons across all 36 tests (six diagnostic groups × six time windows) using the Benjamini-Hochberg false discovery rate procedure. \*q < 0.05 after Benjamini-Hochberg correction. p-values are unadjusted and shown for transparency.

